# Bangla Technique of Laparoscopic Cholecystectomy: a Novel, Safe and Cost-effective modification of American and French Techniques

**DOI:** 10.1101/2020.10.08.20209056

**Authors:** Md Jafrul Hannan, Mosammat Kohinoor Parveen, Alak Kumar Nandy

## Abstract

**Background:** Laparoscopic cholecystectomy (LC) can be performed by following either of the two approaches proposed by the American and the French school. The two approaches have comparable operative times, but use different arrangements for the patient’s and operators’ positions, and sites for port insertions.

The aim of the present paper is to describe an alternative to the American and the French approaches, referred to as the Bangla technique, which uses a standard four port approach but requires the presence of only one assistant along with the surgeon. It is hoped that the Bangla technique will improve surgery outcomes for gallbladder disease patients and encourage healthcare professionals in resource-poor settings to adopt minimally invasive/laparoscopic approaches to surgical problems.

**Methods:** The sample consisted of a total of 280 gallbladder disease retrospective observational cases (of which 21 were children between 6 and 16 of age) who were treated with the Bangla technique at the South Point Hospital Chittagong, Bangladesh, between January 2018 and February 2020.

**Results:** Surgery data showed that using the Bangla technique, the average operating time and average operation theater time were36.25 and 45.9 minutes, respectively. Of the patients, 86% left the hospital on the same day of operation, while the remaining left the following day. In 91.7% of the cases, there were no complications, while content leakage and bleeding occurred in 6.7% and 1.4% cases, respectively.

**Conclusion:** The proposed LC technique will benefit infection prevention and control by reducing the number of personnel in the operation theatre (one assistant and the surgeon) and, as such, reducing surgery-related expenses, which can be further decreased by using only one monitor. More so, the Bangla technique can be combined with the cystic artery sparing technique to reduce the risk of injury to the common bile duct and bleeding.

**Mini Abstract:** Laparoscopic cholecystectomy can be performed following the American or the French approach. The present paper proposes an alternative to the American and the French approaches referred to as the Bangla technique, which uses a standard four port approach but requires the presence of only one assistant along with the surgeon.

## Introduction

Laparoscopic cholecystectomy (LC) is the gold standard for the surgical treatment of gallbladder diseases. The laparoscopic technique can be performed by following either the American [1], or the French approach[2], which are different in terms of positioning of the patients, operators, and trocars. Selecting one technique over the other is usually related to the surgeon’s training and the operator’s habit, as opposed to evidence-based clinical indications[3].

In the American technique, the patient takes a supine position with closed legs while the surgeon places themselves on the patient’s left; one assistant holds the camera to the surgeon’s left, and another assistant places themselves on the patient’s right side. In the French approach, the surgeon stands between the spread legs of the patient and the two assistants are positioned to the patient’s left and to the right side of the surgeon.

The main difference between the two approaches is the port position. In the American approach, one 5 mm port is on the right side of patient at the level of umbilicus while in the French approach this is on the left of the patient’s midline, and above the transverse umbilical line. This difference implies that the traction of the assistant on the bottom of the gallbladder is not exerted from the same position in both techniques. In the American technique, the exertion of traction is done via the right lateral port, and in the French approach, via the epigastric port. The two techniques are similar in terms of the surgical tactics used for traction on the gallbladder, isolation of the elements before clip application, and exposure of the Calot’s triangle. The proponents of each method consider that some maneuvers are simpler in one approach compared to the other, or prefer repeating the same movements in similar phases of an intervention so as to reduce tissue trauma and errors, and to increase the execution speed[3].

While no studies indicate that one of the two methods provides a lower risk of biliary lesions or other major complications, Perissat [4] argues that the use of several traction modes for the gallbladder in the American approach may increase bile duct injury risks. More specifically, in the American technique the liver is retracted by axial traction on the gallbladder through the anterior axillary cannula and the infundibulum through the mid-clavicular cannula. In the French technique, on the other hand, the liver is retracted via the mid-clavicular cannula and the infundibulum of the gallbladder via the anterior axillary port. Moreover, one study showed that the French approach had a lower impact on postoperative lung function [5].

In a pooled data analysis and systematic review analyzing outcome trends and safety measures of LC as reported by 51 studies published in the last three decades, overall morbidity, bile leakage and bile duct injury (BDI), and mortality rates were 1.6-5.3%, 0.32-0.52%, and 0.08-0.14%, respectively [6]. BDI rates reported in 2010-2015 were lowered than those reported in1994-1999 (0.02-0.40% vs 0.52-0.84%, respectively). The meta-analysis indicated a higher conversion rate in developed countries compared to developing countries (4.7% vs 3.4%), although a reporting bias was found in the studies included.

A systematic review of BDI prevention [7] concluded that many studies have used low sample sizes and suboptimal study designs which makes it difficult to indicate whether one method is better compared to others to prevent BDI; however, the paper recommended that surgeons should mainly focus on techniques of proper dissection, of which the Critical View of Safety (CVS) approach[8, 9] is the most suitable. The CVS approach consists of a blunt dissection of the upper part of Calot’s space, which typically does not contain biliary or arterial anomalies and is, as such, is ideal for a safe dissection. The use of CVS is also recommended by the Society of American Gastrointestinal and Endoscopic Surgeons (SAGES) [10].

## Methods

### Description of the Technique

The proposed approach, referred to as the Bangla technique, benefits infection prevention and control by reducing the number of personnel in the operation theatre (one assistant and the operating surgeon) and, as such, reducing surgery-related expenses—which are further reduced by using only one monitor.

A standard four-port procedure is used. The patient is kept in the supine position on the operating table. After anesthesia is administered (in most cases spinal anesthesia), the patient is restrained via a cloth belt at the level of the hips, which prevents them from falling. In our study, the spinal anesthesia consisted of 0.5% Bupivacaine in 8.5% dextrose at a dose of 0.4 mg/kg of body weight. CO2 insufflation pressures were kept under 8 mmHg and flow was maintained between 2.0-2.5 L/minute in children; 12 mmHg and 3.0-3.5 L/minute in adults. Patients also received sedation with intravenous diazepam or ketamine hydrochloride as an adjunct to alleviate their anxiety.

Ports (most often of 10 mm) are then inserted starting with the camera port at the upper or lower fold of the umbilicus. Three additional 5 mm ports are inserted: one in the epigastrium below the xiphisternum, one on the left side at a point on the midclavicular line between the umbilicus and the costal margin, and one on the right side at the level of the umbilicus. The Bangla position can be seen in Figures 1 and 2. Figures 3 and 4 shows how each port is placed on the patient.

**Figure 1.**
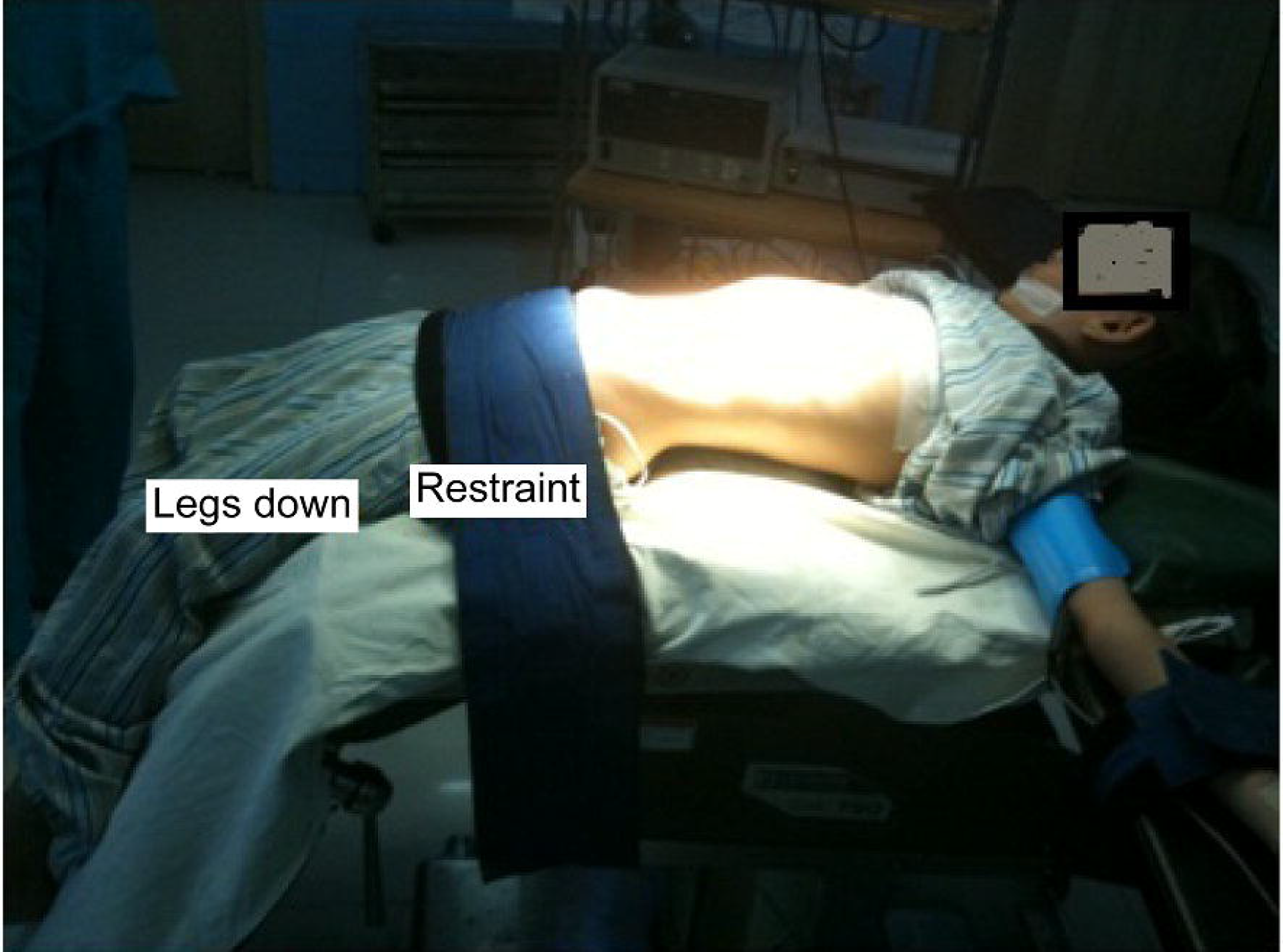
A patient places in the Bangla position before surgery.

**Figure 2.**
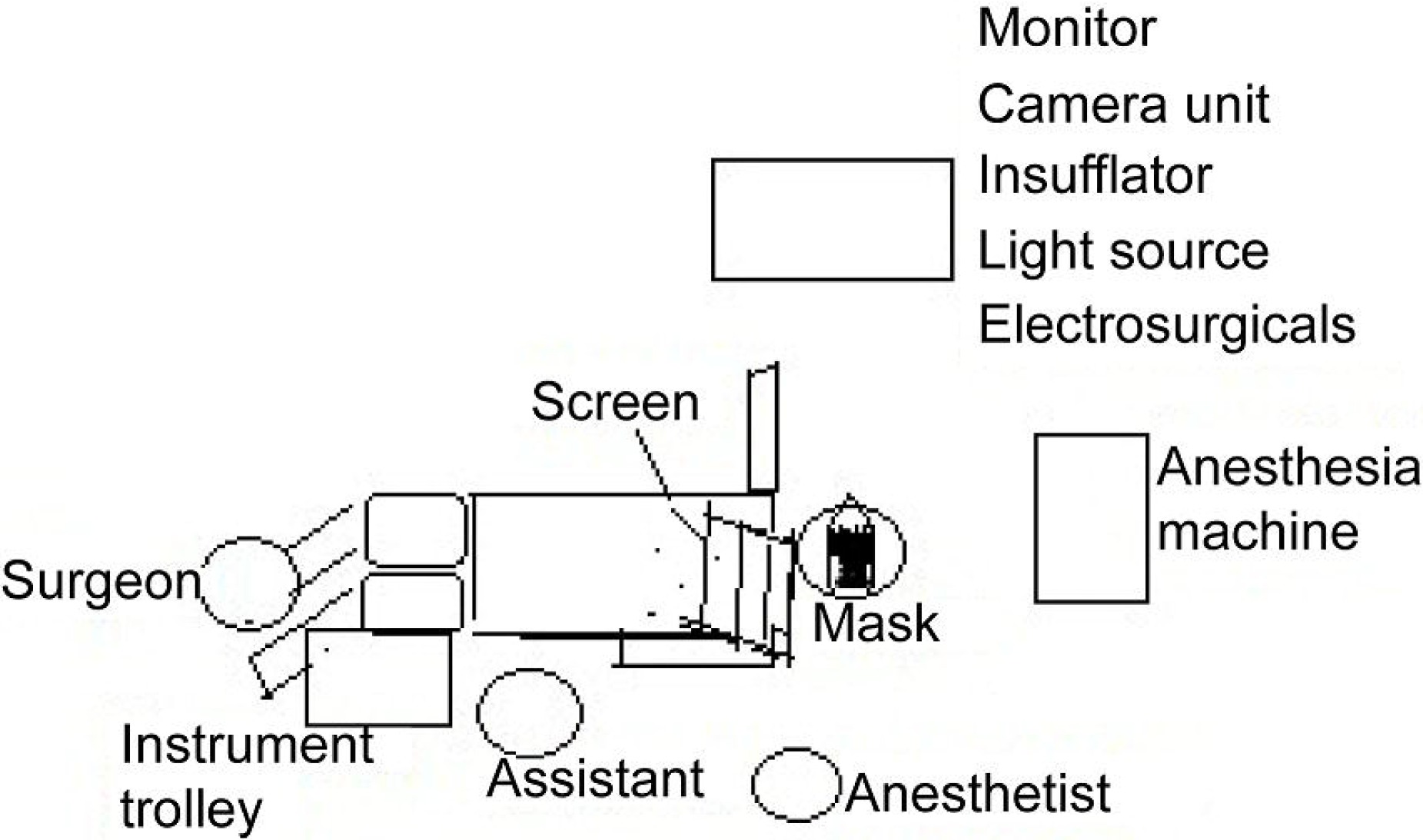
The position of the surgeon, the assistant, the anesthetist, and the instrument trolley in the Bangla position.

**Figure 3.**
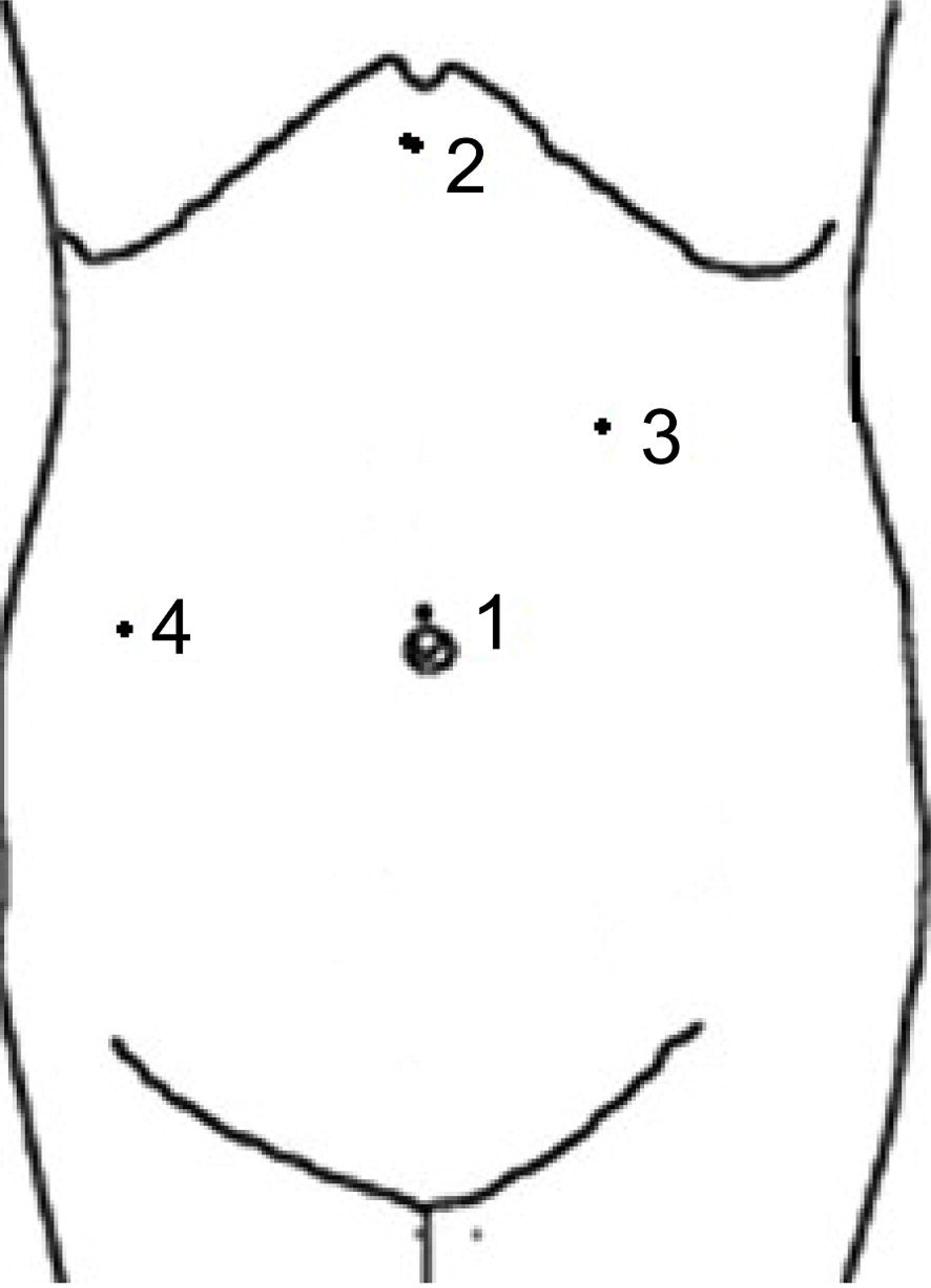
The four ports used in the Bangla position: One at the upper fold of the umbilicus, one in the epigastrium, one on the left side at a point on the midclavicular line and one on the right side at the level of the umbilicus.

**Figure 4.**
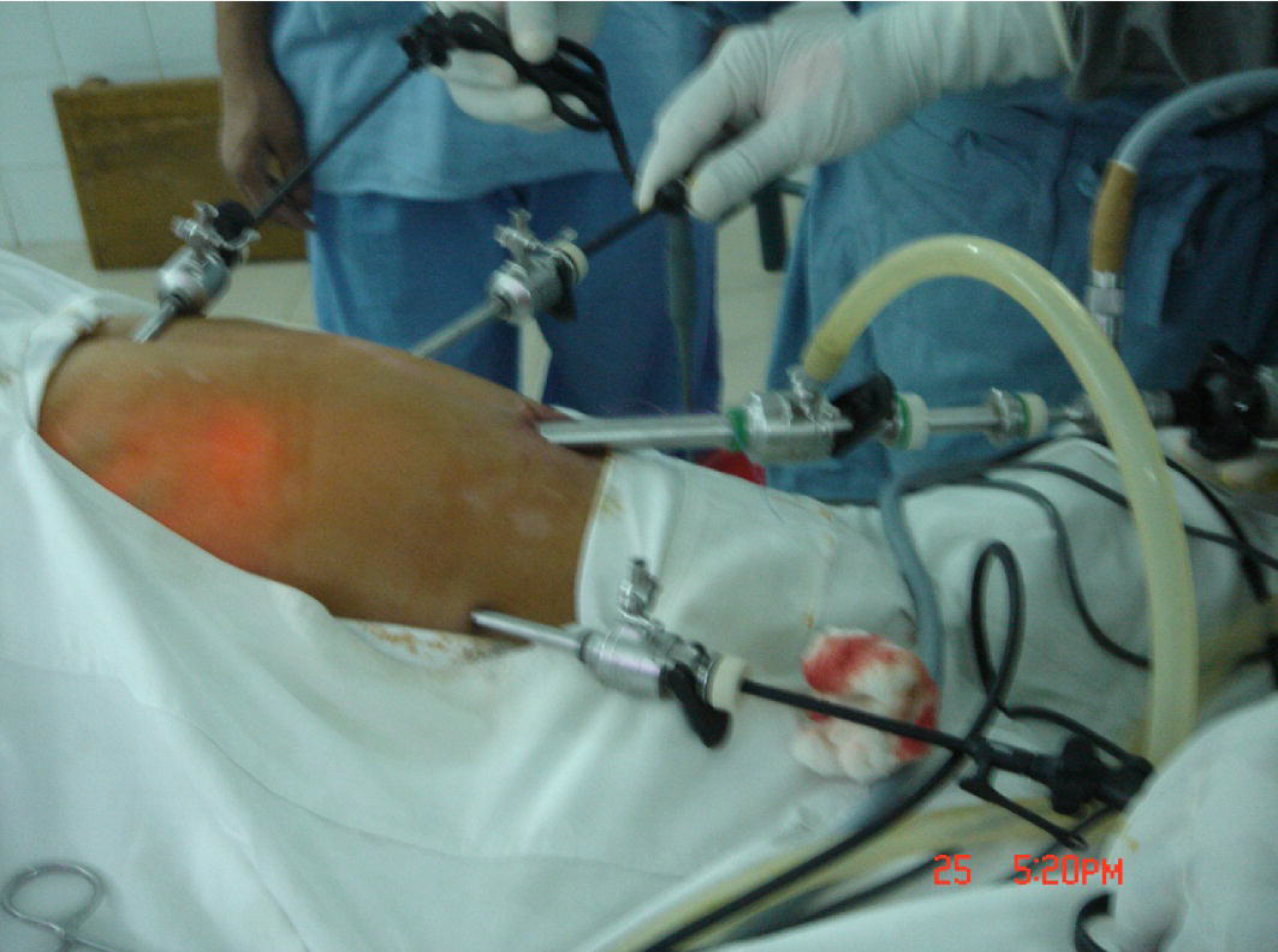
Ports inserted in a patient in the Bangla position.

The foot end of the operating table is lowered along with the patient’s lower limbs. The head end of the operating table is elevated by 25–35 degrees, and the right side of the patient is elevated by 15–20 degrees. The trolley, which includes the monitor, camera unit, light source, insufflator, and electro-surgical units, is placed toward the head end of the patient on the right side. The instrument trolley is kept toward the foot end of the table on the left side of the patient, between the surgeon and assistant.

The surgeon stands in front of the patient’s lower limbs and the assistant stands on the left side of the patient. The surgeon handles the instruments inserted through the left and right ports for the operative steps. The assistant holds the telescopes inserted through the umbilical port with their left hand and holds and pushes away the gall bladder upwards and to the right by using their right hand and the instrument through the epigastric port. Once the gall bladder is freed, it is retrieved through the umbilical port (10 mm) using a reduction sleeve. The visualization during retrieval is realized via the port on the left side with a 5 mm telescope.

To reduce infection risks and complications, the Bangla technique can be combined with the cystic artery sparing technique [11], which allows avoiding handling and injury to the cystic artery and, consequently, ductal injuries. In this technique, dissection starts distal to the cystic lymph node on the gallbladder wall using a monopolar hook cautery or a combination of bipolar and monopolar cautery in many cases; as such, the cystic artery sparing technique does not touch the cystic artery while clearing the Calot’s. The average surgery time was 40 minutes and the surgery outcome data showed no immediate postoperative complications.

### Sampling and Statistical Analysis

The study was undertaken in South Point Hospital Chittagong, Bangladesh between 1st January 2018 and 29th February 2020 (out of which 18 were partial LC cases [12, 13]). The sample consisted of a total of 280 retrospective observational cases who were treated with the Bangla technique. The sample included 21 children between 6 and 16 years of age diagnosed with the gallbladder stone disease and 259 adults up to 60 years of age diagnosed with the gallbladder stone disease, acalculous cholecystitis and gallbladder polyps. Patients of other ages, patients with severe heart disease and/or uncontrolled diabetes, and patients with gallbladder diseases where another physical condition made anesthesia contraindicated where excluded.

Ethical clearance was obtained from the Ethical Committee of South Point Hospital. Consent was obtained from all adult patients and the parent(s) or legal guardians of the minor patients. The study was conducted in accordance to the ethical principles for medical research involving human subjects as provided by the World Medical Association Declaration of Helsinki[14].

## Results

Descriptive statistics were obtained with the IBM Statistical Product and Service Solutions (SPSS) v.27[15]. Data analysis showed an average operating time of 36.25 minutes and an average operation theater time of 45.9 minutes. Of the patients, 86%left the hospital on the same day of operation, while the remaining left the following day. In 91.7% of the cases there were no complications, while content leakage occurred in 6.7% of the cases, and bleeding occurred in 1.4% of the cases.

There were no cases of CBD injury. In children, there were no cases of excessive adhesions of omentum, duodenum, stomach, or intestinal loops obscuring the Calot’s triangle, which would make dissection difficult. Such events are not uncommon in adults; however, cystic artery sparing technique starts dissection relatively proximally on the gall bladder wall, and can locate the Calot’s triangle in a timely manner.

There was no case of open cholecystectomy in children. In adults, during the past 2 years, only two adult cases required opening because of excessive adhesion of the duodenum and stomach to the gall bladder wall, which posed a risk of injury during the use of dissection instruments. Table 1 provides the key statistics for each type of complication.

**Table 1.**
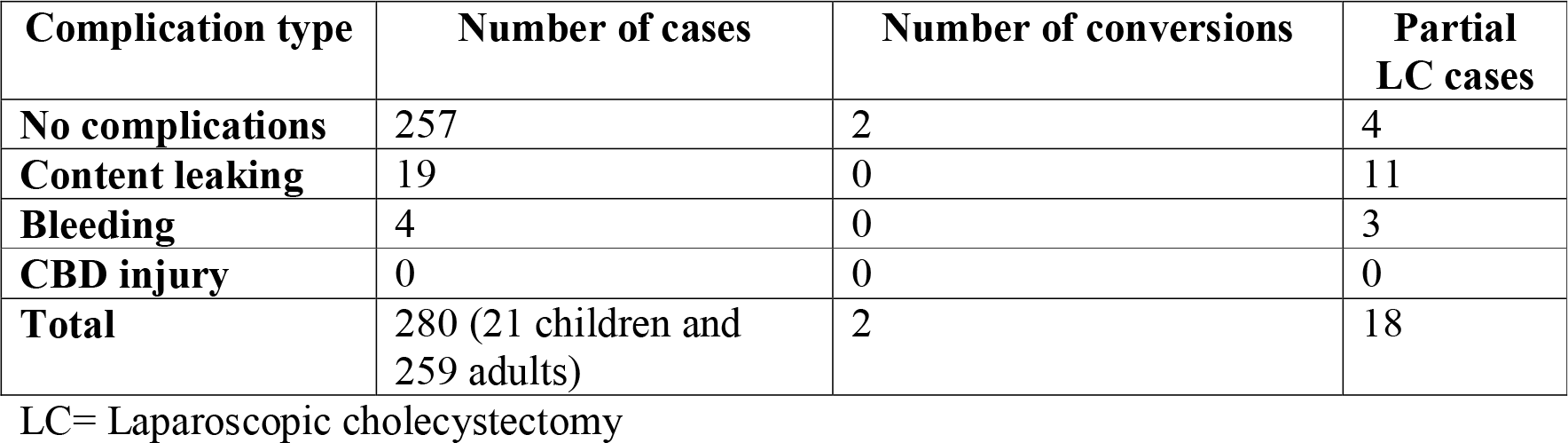
Number of cases, number of conversions, and number of partial laparoscopic cholecystectomy (LC) cases per complication type.

| Complication type | Number of cases | Number of conversions | Partial LC cases |
| --- | --- | --- | --- |
| No complications | 257 | 2 | 4 |
| Content leaking | 19 | 0 | 11 |
| Bleeding | 4 | 0 | 3 |
| CBD injury | 0 | 0 | 0 |
| Total | 280 (21 children and 259 adults) | 2 | 18 |

LC= Laparoscopic cholecystectomy

## Discussion

The present paper proposes an alternative to the American and the French approaches, referred to as the Bangla technique, which uses a standard four-port approach but requires the presence of only one assistant along with the surgeon. The American technique requires two assistants and one scrub nurse besides the surgeon. The French technique can be performed with one less assistant, while the Bangla technique eliminates the need for a scrub nurse in the operative team. Based on the evidence described, we argue that complications are not related to the technique (American/French/Bangla), but to the level of expertise of the surgeon and the inherent condition of the gall bladder and adjacent structures. On the other hand, the dissection technique used during the operation matters. As mentioned in an earlier section, the CVS and the cystic artery sparing technique described reduces complications and conversion to an open procedure.

All laparoscopic surgeries performed using the Bangla technique, including in children above 5 years of age, were conducted under spinal anesthesia. Regional anesthesia, such as spinal, epidural, and combined spinal-epidural anesthesia come with several advantages over general anesthesia, including an awake and spontaneously breathing patient intraoperatively, the prevention of airway manipulation, effective post-operative analgesia, minimal nausea and vomiting, and early ambulation and recovery; neverhteless, side effects such as ventilatory changes due to requirement of a higher sensory level, more severe hypotension, respiratory embarrassment caused by the pneumoperitoneum, and shoulder discomfort due to diaphragmatic irritation should also be considered[16]. On the other hand, in children, the hemodynamic response appears to be insignificant in the case of spinal anesthesia [17], or less significant than in general anesthesia[18]. More so, unlike what often happens during general anesthesia, ventilatory support for respiratory depression does not seem to be required, stress hormones are not released, and postoperative analgesia is not necessary. Spinal anesthesia also saves a considerable amount of operation theater time, about 15-30 minutes, as it obviates the need for induction and reversal of anesthesia[18].

Reducing the number of operative team personnel, as with the Bangla technique, will encourage healthcare professionals and administrators in resource-poor settings with shortages of skilled manpower to adopt minimally invasive/laparoscopic approaches to surgical problems. The Bangla technique combined with the cystic artery sparing technique, with spinal anesthesia use, provides a suitable option for a safe, cost-effective LC that poses lesser infection risks.

It is hoped that the Bangla technique will be implemented in more hospitals across Bangladesh and in other parts of Asia and the world, which will allow future studies to replicate the current study with a much larger sample; hence providing data with a high degree of validity and generalizability.

## Data Availability

All data regarding this manuscript will be free to access

## Acknowledgements

**None**

